# Exploration of interethnic variation and repurposed drug efficacy in the treatment of SARS-CoV-2 Infection (COVID-19)

**DOI:** 10.1101/2021.03.07.21253095

**Authors:** Ammar Ali Almarzooq

## Abstract

The COVID-19 global pandemic has led to repurposing of drugs, with little underlying evidence for treatment safety and efficacy. This may increase complications for patients with acute viral respiratory infections. *UGT1A1* and *CYP2D6* enzymes are involved in the metabolism of atazanavir and fluvoxamine repurposed for COVID-19. This study aimed to elucidate the role of interethnic variation in these enzymes in the efficacy of repurposed drug therapies. A retrospective cohort of 101 Jordanian Arab samples were genotyped using Affymetrix DMET Plus Premier Package. Comprehensive global population genetic structure analyses were performed for *CYP2D6* and *UGT1A1* allele frequencies across multi-ethnic populations of over 131,000 global subjects from 417 published reports, revealing that Jordanian Arabs share the closest sequence homology to European and Near East populations. The East Asian populations have significantly over-representaiton of individuals with diplotype pairs for reduced atazanavir metabolism compared to the African populations and are more likely to show impaired UGT1A1 metabolism. East Asian populations are also 4.4x more likely to show impaired fluvoxamine metabolism than South Central Asian and Oceanian populations, and 8x more likely than other ancestry populations. The results here support previous findings that interethnic variation should be used for developing proper population-specific dosage guidelines.

## INTRODUCTION

The global coronavirus disease 2019 (COVID-19) pandemic, caused by the severe acute respiratory syndrome coronavirus 2 (SARS-CoV-2), has led to more than 105 million infected patients and more than 2,600,000 deaths worldwide^1^. With the rise of multiple and potentially more virulent variant strains, managing the treatment and spread of the disease is paramount. The management of the disease includes preventive and therapeutic strategies, treatment of the acute respiratory distress syndrome and the cytokine storm^2^. Patients who are hospitalized with COVID-19 may be treated with multiple therapeutics, including repurposed treatments and adjuvant therapies^3,4^, in addition to any concomitant medications already prescribed to the patients^5^. As such, it is vital for clinicians to be able to quickly and easily integrate patient specific factors, such as genetic variation to optimize treatment^6,7^.

Many of the investigational repurposed drugs have clinical pharmacogenetic guidelines available, with therapeutic recommendations indicated on the drug label^8^. Atazanavir and fluvoxami,ne for example, include both pharmacogenetics information and prescription recommendations. A molecular dynamics analysis and *in vitro* experiments showed that atazanavir, a protease inhibitor against human immunodefiency virus (HIV), could inhibit SARS-CoV-2 replication, alone or in combination with ritonavir^9^. Atazanavir inhibits the hepatic uridine diphosphate glucuronosyltransferase family 1 member A1 (UGT1A1), thereby preventing the glucuronidation and elimination of bilirubin^10^. However, resultant indirect hyperbilirubinemia with jaundice, can untimely cause premature discontinuation of atazanavir^11^. Genome-wide association studies (GWAS) that genotyped single nucleotide polymorphisms (SNPs) have consistently associated the polymorphism rs887829 (c.-364C>T; *UGT1A1*80*) with indirect hyperbilirubinemia in the general population (i.e. Gilbert syndrome)^12-14^. The CPIC dosing guideline on atazanavir and *UGT1A1* classifies individuals according to *UGT1A1* alleles into normal metabolizers (NMs), intermediate metabolizers (IMs) and poor metabolizers (PMs). The PMs (i.e. carriers of two decreased function alleles) are more likely to develop jaundice, which may cause non-adherence to medications, and alternative drugs need to be considered. The risk of discontinuation is low or very low for individuals carrying one or zero decreased-function alleles (IMs and NMs)^15^. Meanwhile, the recent report of Lenze and colleagues indicated that the selective serotonin reuptake inhibitor (SSRI) fluvoxamine might reduce the rate of clinical deterioration in outpatients with symptomatic COVID-19^16^. Fluvoxamine, an antidepressant, is primarily used for the treatment of obsessive–compulsive disorder, but it is also used for depression and anxiety disorders, such as panic disorder, social anxiety disorder and post-traumatic stress disorder^17^. Fluvoxamine has been considered for use in the early symptom treatment of COVID-19. Although there are no recommendations for the treatment of COVID-19 with fluvoxamine, a Phase I clinical trial is currently being conducted on outpatients to assess if serious complications, such as shortness of breath, can be prevented^18^. It was previously -shown that fluvoxamine reduced the inflammatory response “cytokine storm” during sepsis^19^. The second phase of COVID 19 can involve a serious inflammatory reaction^20^, which may be prevented if fluvoxamine is used for treatment. However, SSRIs including fluvoxamine increase the risk of developing gastrointestinal side effects when a non-efficient cytochrome P450 (*CYP2D6*) enzyme variant is present^21^. When administered in similar doses, *CYP2D6* poor metabolizers have significantly greater exposure to fluvoxamine when compared to NMs^21,22^. This increase in drug exposure may be a risk factor for drug-induced side effects or toxicity. The US Food and Drug Administration (FDA) states that fluvoxamine should be used cautiously in patients known to have reduced levels of CYP2D6 activity^23^. The Clinical Pharmacogenetics Implementation Consortium (CPIC) guideline recommends a 25–50% dose reduction for the starting dose of fluvoxamine, or the use of an alternative drug that is not metabolized by enzyme CYP2D6 for *CYP2D6* PMs (*\*3/*4, *4/*4, *5/*5*, and *\*5/*6*)^24^. Therefore, host genetics and demography associated with COVID-19 are crucial aspects of infection and prognosis. Hence, integral medication dosing might need adjustments based on a patient’s genetic information.

The CPIC has published a pharmacogenetic guideline on SSRI and atazanavir, with specific therapeutic recommendations based on *CYP2D6* and *UGT1A1* phenotypes, respectively^15,24^. The phenotype was derived from an activity score, obtained by the sum of two individuals’ allele scores. However, recent meta-analysis of concentrations using the Pharmacogenomics Knowledge Base (PharmGKB), showed strong correlations between the *UGT1A1*80* allele with higher plasma levels of atazanavir, and the *CYP2D6*3, *4, *5, *6* and *\*10* alleles with higher plasma levels of fluvoxamine^25^. Based on these results, PharmGKB assigned the highest level of evidence (level 1A) to these associations, indicating that these biomarkers were extensively studied for years and are already validated in clinical practice.

The goal of this study was to demonstrate that current pharmacogenomics databases can be leveraged to enhance the identification of gene alleles, and to determine population differences in associated drug response/toxicity events. Results from this work have wide ranging impacts on the targeted treatment of COVID-19 patients across broad geographic ranges and ethnic backgrounds, thereby facilitating the drug development processes and guide towards safer therapeutic prescriptions.26272829

## RESULTS

### Selection and analysis of UGT1A1 and CYP2D6 Variations

Sixty-two variants across 101 Jordanian individuals of Arab descent associated with reduced *UGT1A1* and *CYP2D6* enzymes function were selected (Table 1). Allele and genotype frequencies of the selected variants are listed in Table 2. SNPs were tested for Hardy– Weinberg Equilibrium (HWE), and all the studied polymorphisms did not deviate from the HWE (p-value > 0.05). For *CYP2D6*, the defective allele *\*2* was the most abundant variant (0.243), followed by allele *\*4* (0.163), while the defective allele *UGT1A1*80* was the most abundant variant (0.381), followed by allele *\*28* (0.371). In addition, three less common alleles, *CYP2D6 *10, *17* and *\*41* were also detected. *UGT1A1* and *CYP2D6* common alleles together accounted for 0.752 and 0.619, respectively.

**Table 1.** Details of the 62 allele frequencies of *CYP2D6* and *UGT1A1* genes.

| rsID | Common Name/HGVS | Allele | Call<br>Rate | MAF |
| --- | --- | --- | --- | --- |
| rs72551349 | UGT1A1*10_c.1021C>T (R341X) | *10 | 99 | 0.000 |
| rs62625011 | UGT1A1*11_c.923G>A (G308E) | *11 | 100 | 0.000 |
| rs3755319 | UGT1A1*112_c.-1353A>C | *112 | 99 | 0.545 |
| rs72551341 | UGT1A1*12_c.524T>A (L175Q) | *12 | 100 | 0.000 |
| rs72551345 | UGT1A1*14_c.826G>C (G276R) | *14 | 100 | 0.000 |
| rs72551342 | UGT1A1*15_c.529T>C (C177R) | *15 | 100 | 0.000 |
| rs72551351 | UGT1A1*16_c.1070A>G (Q357R) | *16 | 100 | 0.000 |
| rs72551354 | UGT1A1*17_c.1143C>G (S381R) | *17 | 99 | 0.000 |
| rs72551355 | UGT1A1*18_c.1201G>C (A401P) | *18 | 99 | 0.000 |
| rs72551352 | UGT1A1*20_c.1102G>A (A368T) | *20 | 100 | 0.000 |
| rs72551357 | UGT1A1*24_c.1309A>T (K437X) | *24 | 100 | 0.000 |
| rs35350960 | UGT1A1*27_c.686C>A (P229Q) | *27 | 100 | 0.000 |
| rs3064744 | UGT1A1*28_c.TATA-box (Promoter) | *28 | 99 | 0.371 |
| rs55750087 | UGT1A1*29_c.1099C>G (R367G) | *29 | 100 | 0.000 |
| rs72551353 | UGT1A1*3_c.1124C>T (S375F) | *3 | 100 | 0.000 |
| rs72551350 | UGT1A1*4_c.1069C>T (Q357X) | *4 | 100 | 0.000 |
| rs72551344 | UGT1A1*43_c.698T>G (L233R) | *43 | 100 | 0.000 |
| rs72551340 | UGT1A1*45_c.222C>A (Y74X) | *45 | 100 | 0.000 |
| rs72551361 | UGT1A1*55_c.1487T>A (L496X) | *55 | 99 | 0.000 |
| rs4148323 | UGT1A1*6_c.211G>A (G71R) | *6 | 100 | 0.000 |
| rs4124874 | UGT1A1*60_c.-3279T>G (Promoter) | *60 | 100 | 0.564 |
| rs56059937 | UGT1A1*62_c.247T>C (F83L) | *62 | 100 | 0.000 |
| rs10929303 | UGT1A1*76_c.*211C>T (3'UTR) | *76 | 99 | 0.238 |
| rs1042640 | UGT1A1*78_c.*339C>G (3'UTR) | *78 | 99 | 0.198 |
| rs8330 | UGT1A1*79_c.*440C>G (3'UTR) | *79 | 99 | 0.223 |
| rs72551343 | UGT1A1*8_c.625C>T (R209W) | *8 | 99 | 0.000 |
| rs887829 | UGT1A1*80_c.-364C>T | *80 | 100 | 0.381 |
| rs72551348 | UGT1A1*9_c.992A>G (Q331R) | *9 | 100 | 0.000 |
| rs10929302 | UGT1A1*93_c.-3156G>A (Promoter) | *93 | 99 | 0.361 |
| rs28900406 | UGT1A1_c.1428C>T (P476P) |  | 99 | 0.000 |
| rs1042709 | UGT1A1_c.1531G>C (A511P) |  | 100 | 0.000 |
| rs1976391 | UGT1A1_c.-2950A>G |  | 99 | 0.381 |
| rs5030863 | CYP2D6*11_883G>C (SpliceDefect) | *11 | 100 | 0.000 |
| rs5030862 | CYP2D6*12_124G>A (G42R) | *12 | 100 | 0.000 |
| rs5030865 | CYP2D6*14or*8_1758G>A>T (G169R/X) | *14/*8 | 100 | 0.000 |
| rs72549357 | CYP2D6*15_137insT | *15 | 100 | 0.000 |
| rs1135836 | CYP2D6*18_4125dupGTGCCCACT | *18 | 100 | 0.000 |
| rs72549353 | CYP2D6*19_2539delAACT | *19 | 99 | 0.000 |
| rs72549354 | CYP2D6*20_1973insG | *20 | 100 | 0.000 |
| rs72549352 | CYP2D6*21_2573insC | *21 | 100 | 0.000 |
| rs61736512 | CYP2D6*29_1659G>A (V136I) | *29 | 100 | 0.015 |
| rs59421388 | CYP2D6*29_3183G>A (V338M) | *29 | 99 | 0.010 |
| rs35742686 | CYP2D6*3_2549delA (R259X) | *3 | 100 | 0.015 |
| rs72549351 | CYP2D6*38_2587delGACT | *38 | 100 | 0.000 |
| rs3892097 | CYP2D6*4_1846G>A (SpliceDefect) | *4 | 99 | 0.163 |
| rs72549356 | CYP2D6*40_1863ins (TTTCGCCCC)2 | *40 | 99 | 0.000 |
| rs28371725 | CYP2D6*41_2988G>A (SpliceDefect) | *41 | 99 | 0.149 |
| rs72549346 | CYP2D6*42_3259insGT | *42 | 99 | 0.000 |
| rs72549349 | CYP2D6*44_2950G>C (SpliceDefect) | *44 | 100 | 0.000 |
| rs72549347 | CYP2D6*56_3201C>T (R344X) | *56 | 99 | 0.000 |
| rs5030655 | CYP2D6*6_1707delT (W152X) | *6 | 100 | 0.059 |
| rs5030867 | CYP2D6*7_2935A>C (H324P) | *7 | 100 | 0.005 |
| rs28371720 | CYP2D6*9_2615delAAG | *9 | 99 | 0.000 |
| rs1065852 | CYP2D6_100C>T (P34S) |  | 100 | 0.193 |
| rs28371706 | CYP2D6_1023C>T (T107I) |  | 99 | 0.020 |
| rs1080985 | CYP2D6_-1584C>G |  | 100 | 0.832 |
| rs1058164 | CYP2D6_1661G>C (V136V) |  | 99 | 0.673 |
| rs1080983 | CYP2D6_-1770G>A |  | 99 | 0.728 |
| rs1601813085 | CYP2D6_-1961C>G>A |  | 100 | 0.000 |
| rs28360521 | CYP2D6_-2178G>A |  | 100 | 0.238 |
| rs16947 | CYP2D6_2850C>T (R296C) |  | 100 | 0.441 |
| rs1135840 | CYP2D6_4180G>C (S486T) |  | 100 | 0.619 |

MAF = minor allele frequency

**Table 2.**
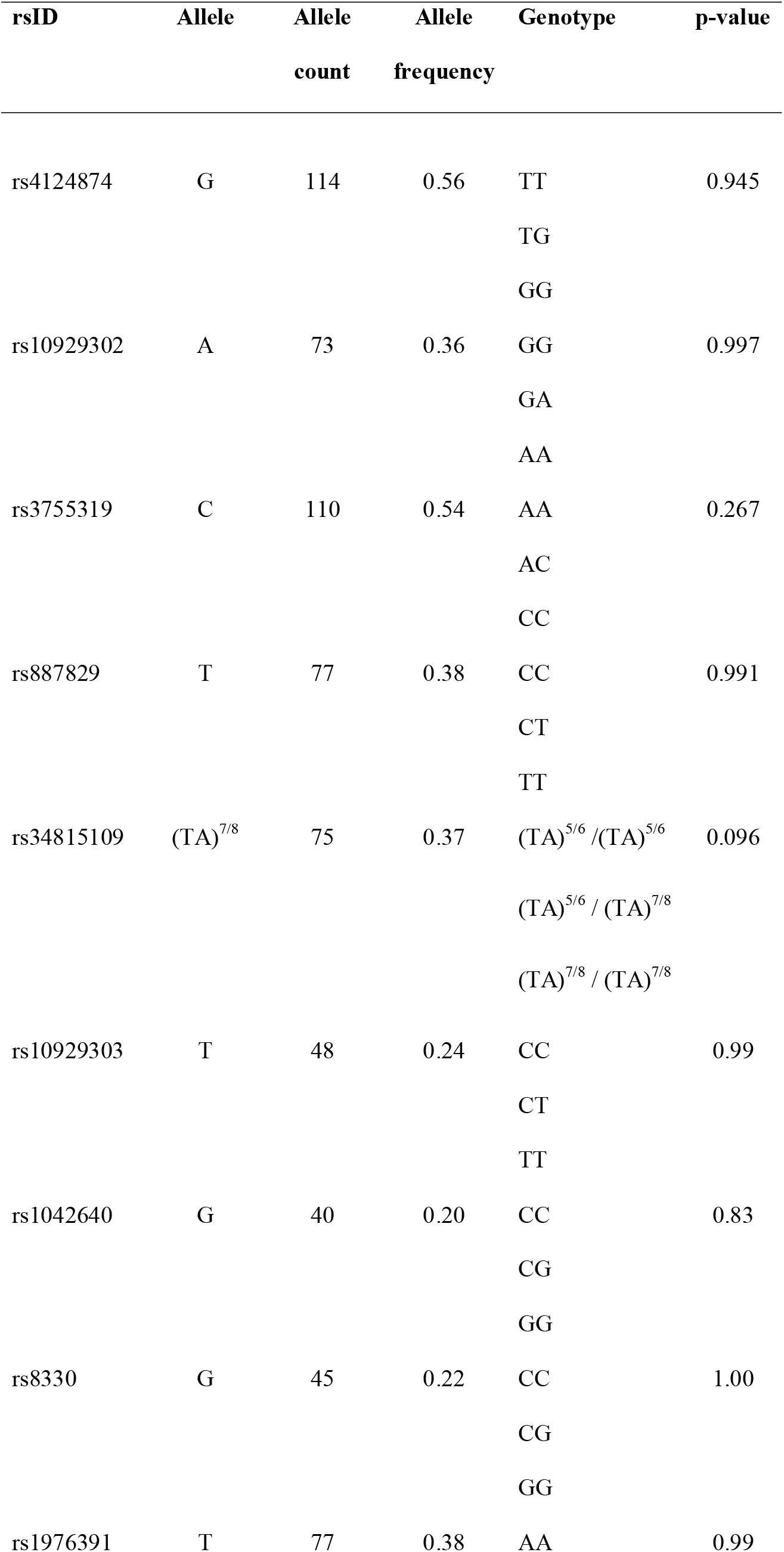
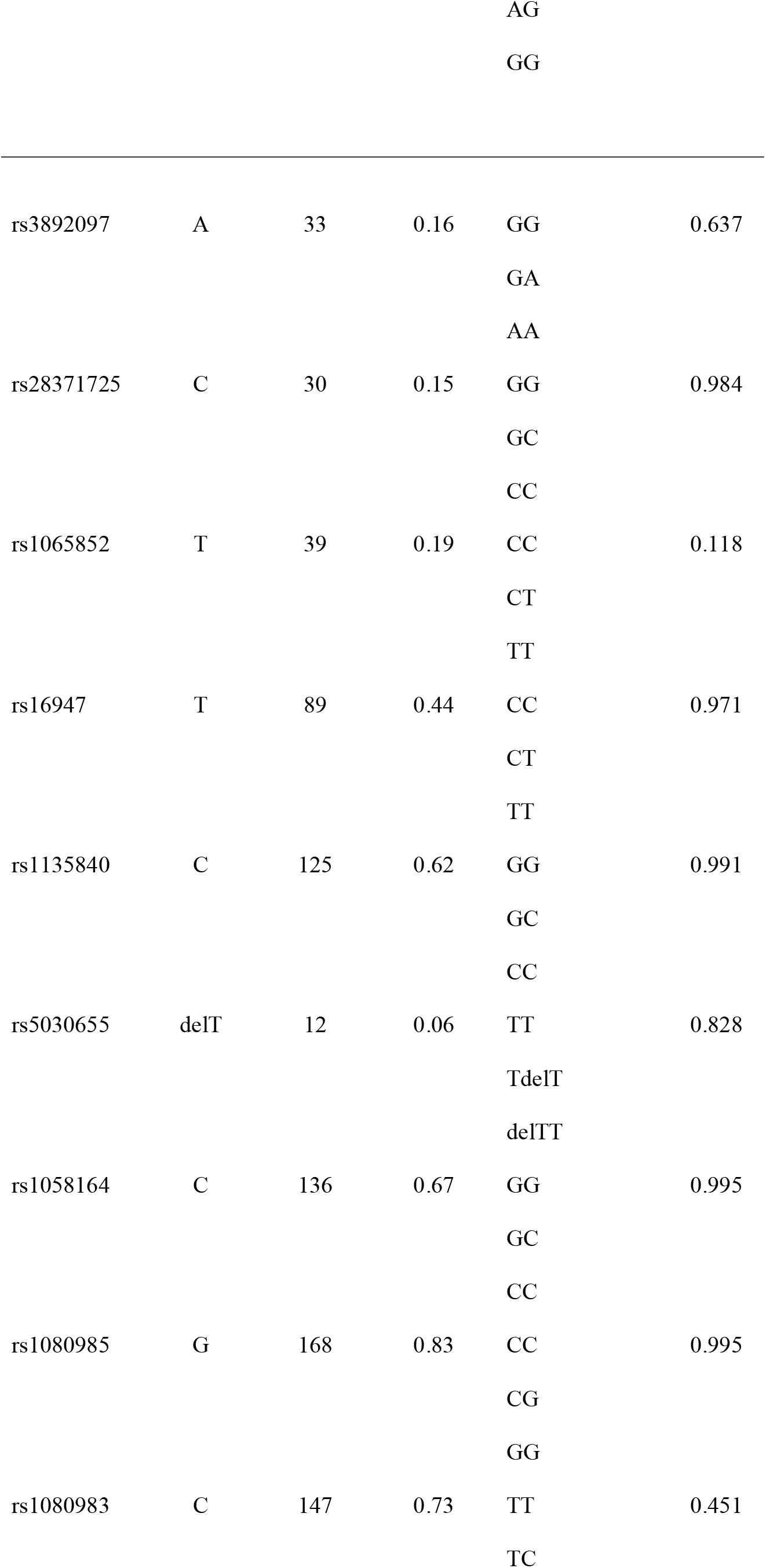
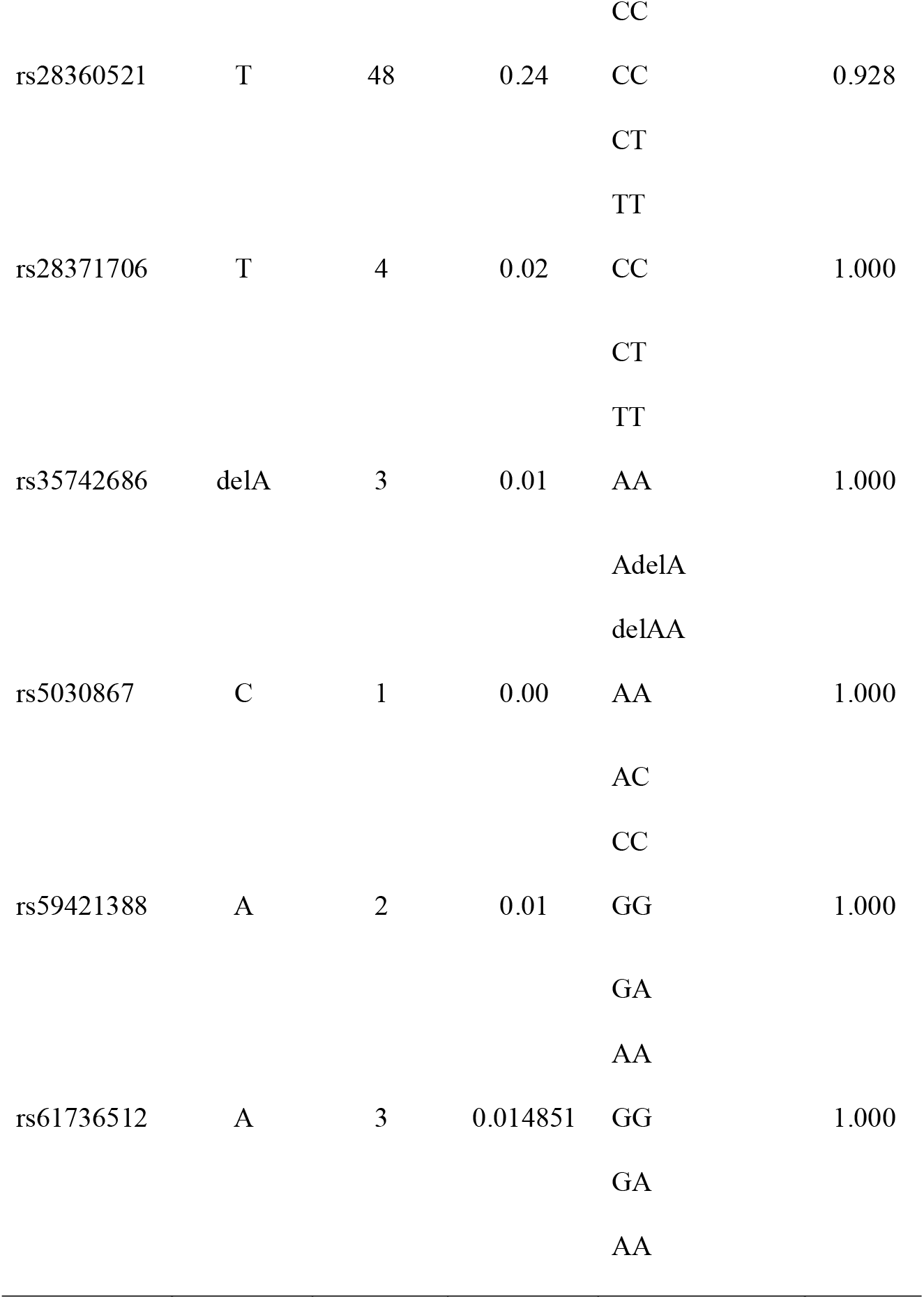
Distribution of alleles and genotypes in Jordanian Arabs across *UGT1A1* and *CYP2D6* genes

| rsID | Allele | Allele<br>count | Allele<br>frequency | Genotype | p-value |
| --- | --- | --- | --- | --- | --- |
| rs4124874 | G | 114 | 0.56 | TT<br>TG<br>GG | 0.945 |
| rs10929302 | A | 73 | 0.36 | GG<br>GA<br>AA | 0.997 |
| rs3755319 | C | 110 | 0.54 | AA<br>AC<br>CC | 0.267 |
| rs887829 | T | 77 | 0.38 | CC<br>CT<br>TT | 0.991 |
| rs34815109 | (TA)<sup>7/8</sup> | 75 | 0.37 | (TA)<sup>5/6</sup> / (TA)<sup>5/6</sup><br>(TA)<sup>5/6</sup> / (TA)<sup>7/8</sup><br>(TA)<sup>7/8</sup> / (TA)<sup>7/8</sup> | 0.096 |
| rs10929303 | T | 48 | 0.24 | CC<br>CT<br>TT | 0.99 |
| rs1042640 | G | 40 | 0.20 | CC<br>CG<br>GG | 0.83 |
| rs8330 | G | 45 | 0.22 | CC<br>CG<br>GG | 1.00 |
| rs1976391 | T | 77 | 0.38 | AA | 0.99 |
|  |  |  |  | AG |  |
|  |  |  |  | GG |  |
| rs3892097 | A | 33 | 0.16 | GG | 0.637 |
|  |  |  |  | GA |  |
|  |  |  |  | AA |  |
| rs28371725 | C | 30 | 0.15 | GG | 0.984 |
|  |  |  |  | GC |  |
|  |  |  |  | CC |  |
| rs1065852 | T | 39 | 0.19 | CC | 0.118 |
|  |  |  |  | CT |  |
|  |  |  |  | TT |  |
| rs16947 | T | 89 | 0.44 | CC | 0.971 |
|  |  |  |  | CT |  |
|  |  |  |  | TT |  |
| rs1135840 | C | 125 | 0.62 | GG | 0.991 |
|  |  |  |  | GC |  |
|  |  |  |  | CC |  |
| rs5030655 | delT | 12 | 0.06 | TT | 0.828 |
|  |  |  |  | TdelT |  |
|  |  |  |  | delTT |  |
| rs1058164 | C | 136 | 0.67 | GG | 0.995 |
|  |  |  |  | GC |  |
|  |  |  |  | CC |  |
| rs1080985 | G | 168 | 0.83 | CC | 0.995 |
|  |  |  |  | CG |  |
|  |  |  |  | GG |  |
| rs1080983 | C | 147 | 0.73 | TT | 0.451 |
|  |  |  |  | TC |  |
|  |  |  |  | CC |  |
| rs28360521 | T | 48 | 0.24 | CC | 0.928 |
|  |  |  |  | CT |  |
|  |  |  |  | TT |  |
| rs28371706 | T | 4 | 0.02 | CC | 1.000 |
|  |  |  |  | CT |  |
|  |  |  |  | TT |  |
| rs35742686 | delA | 3 | 0.01 | AA | 1.000 |
|  |  |  |  | AdelA |  |
|  |  |  |  | delAA |  |
| rs5030867 | C | 1 | 0.00 | AA | 1.000 |
|  |  |  |  | AC |  |
|  |  |  |  | CC |  |
| rs59421388 | A | 2 | 0.01 | GG | 1.000 |
|  |  |  |  | GA |  |
|  |  |  |  | AA |  |
| rs61736512 | A | 3 | 0.014851 | GG | 1.000 |
|  |  |  |  | GA |  |
|  |  |  |  | AA |  |

### Genetic structure of UGT1A1 and CYP2D6 across populations

Genetic structure analysis across populations validated our findings that Jordanian Arabs share the closest variants sequence homology to Near East and European populations (Fig. 1) The two leading principal components from the 30 variants shared between the Jordanian Arab population and the 22 global populations from the 1000 Genomes Project Phase III (1kG-p3) dataset, showed a well-defined separation between the Jordanian Arab population and East Asian and African super populations (Fig. 1a).

**Figure 1.**
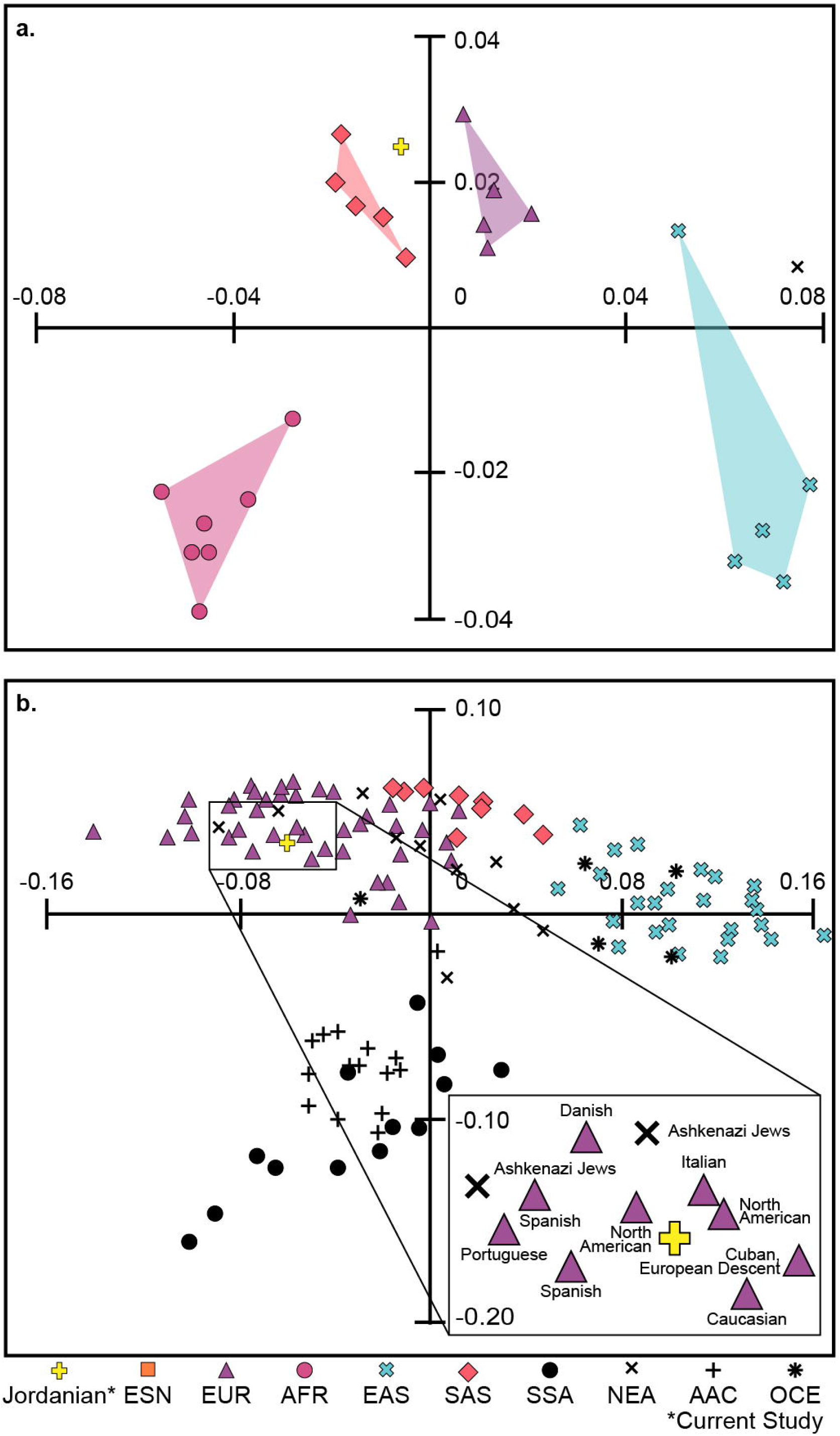
Population structure analyses. (**a**) MDS plot of the 30 variants shared between the Jordanian Arab population and 22 global populations from the 1000 Genomes Project Phase III (1kG-p3) dataset. (**b**) MDS plot of the Jordanian Arab population and 121 public worldwide populations.

Lack of genomic data for additional ethnic groups in the 1000 Genomes Project, such as Near Eastern populations, can reduce robustness and potentially result in biased geographic-based genomic analysis. Therefore, a secondary analysis was performed to include under-represented populations. The two leading principal components shared between the Jordanian Arab population and the 121 global reports including the Near Eastern population, for *CYP2D6*4* and *\*17* suggested a well-defined genetic separation between Jordanian Arabs and East Asian and African super populations, as well as with Oceanian and Central or South Asian populations (Fig. 1b). In addition, a defined cluster of European and Near Eastern populations were found, which was further validated using pairwise Fst analyses (Table S1). The lowest level of differentiation was observed between the Jordanian Arab population and a North American population which was self-defined as ‘Caucasian’ and living in the United States (Fst = 1.48×10^−5^), followed by Portuguese (Fst = 2.88×10^−5^) and Iberian from Portugese and Spanish populations (Fst = 1.34×10^−4^). The greatest affinity with Near Eastern was observed with the Ashkenazi Jews in Argentina (Fst = 6.75×10^−4^).

### Pharmacogenetic analyses by biogeographic grouping system

Across the nine biogeographical groups, about 50% of subjects were of Europeans origin, followed by East Asian (17%), African Americans/Afro-Caribbean’s (10%), Latinos (8%), Sub-Saharan Africans (4%), South Central Asians (4%), Americans (4%), Near Easterns (3%), and Oceanians (1%; Table 3). Distinct differences were found among these populations, with direct impacts on atazanavir and fluvoxamine clinical outcomes (Fig. 2). The *UGT1A1*80* allele frequency was significantly higher in the Sub-Saharan African origin (0.209), followed by African American/Afro-Caribbeans (0.208), South Central Asians (0.184), Jordanian Arabs (0.149), Latinos (0.138) and Europeans (0.087), indicating a decreased metabolism and clearance of atazanavir as compared to East Asians (0.03; Table S2). These significant variant alleles and genotypes were classified as PharmGKB Level 1A evidence with reduced enzyme function, and are therefore associated with recommended changes to atazanavir dosing^15^. Inferring from the specific dosing guidelines for individual with diplotype pairs for reduced fluvoxamine metabolism^24^, the results here indicates that the East Asian populations are about 8x more likely to show impaired *CYP2D6* metabolism than African, European, Near Eastern, American and Latino populations, and 4.4x more likely than South Central Asian and Oceanian populations (Table S2).

**Table 3.**
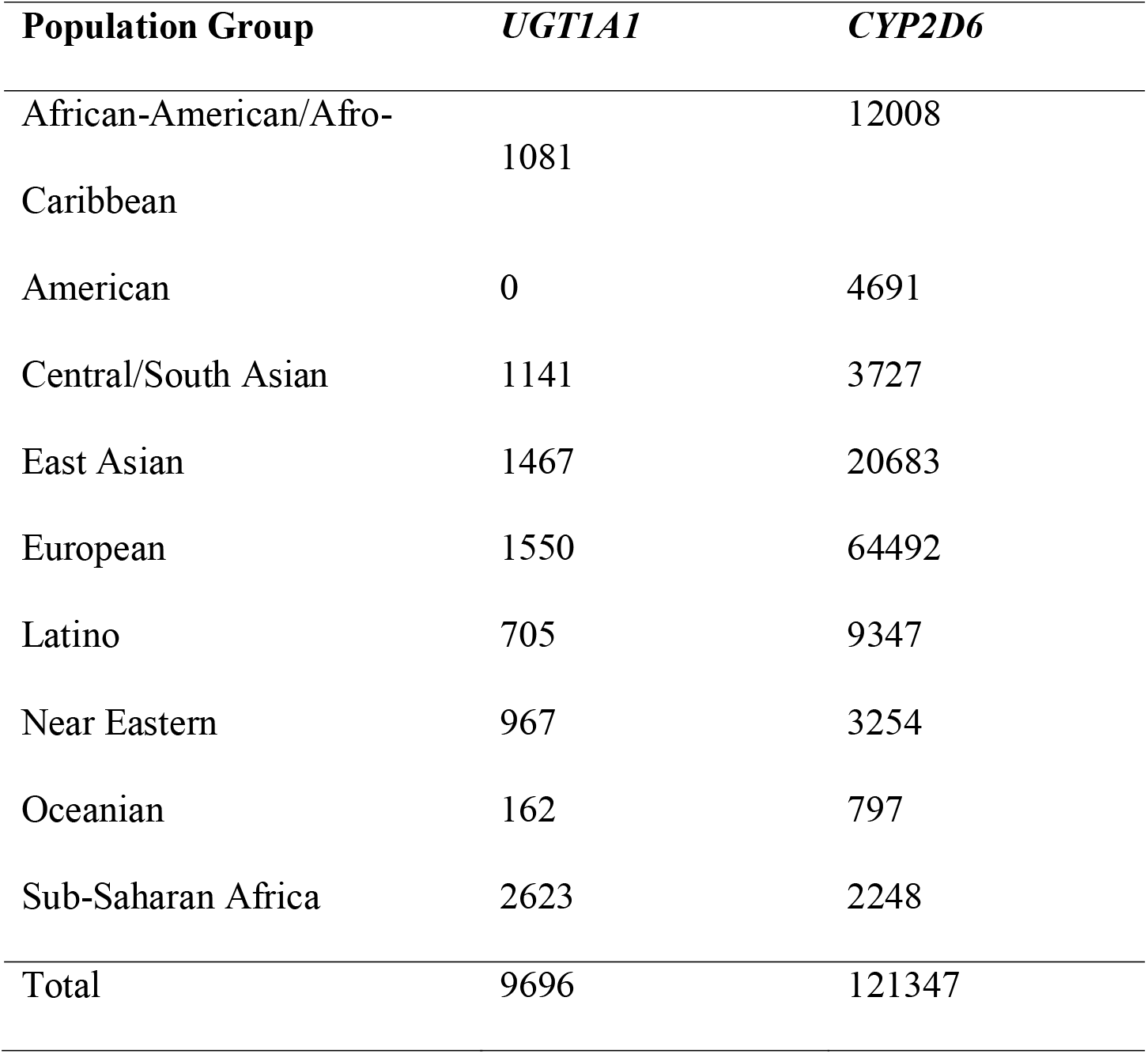
Sample size accumulated per biogeographical group.

**Figure 2.**
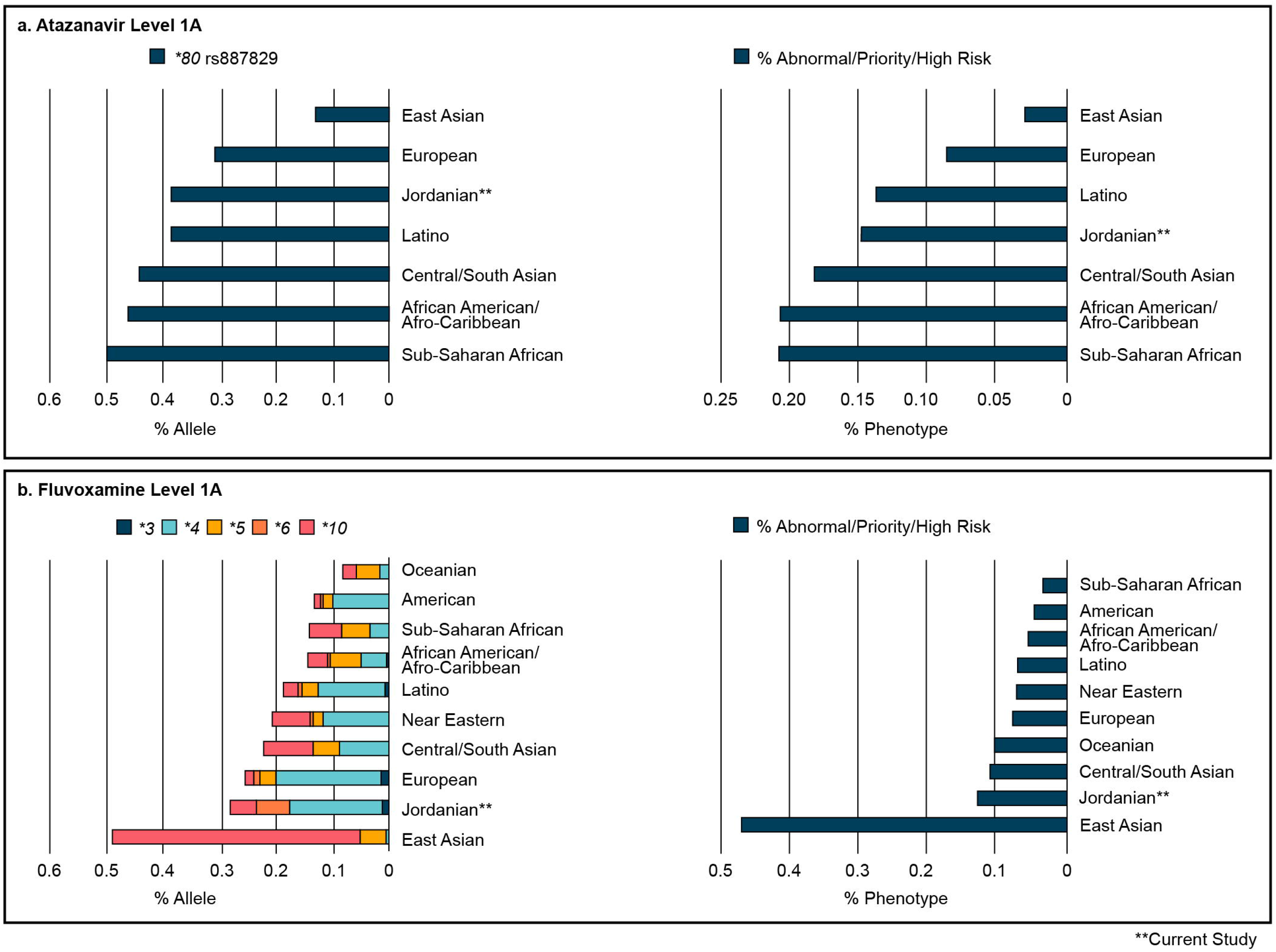
Biogeographical groups and pharmacogenetic analyses of the total allele and phenotype frequency, strongly associated with PharmGKB’s level 1A. (**a**) Mapping of the total allele and phenotype frequency of the *UGT1A1* allele *80 (defined by rs887829). (**b**) Mapping of the total allele and phenotype frequency of the *CYP2D6* alleles *3, *4, *5, *6 and *10.

Interestingly, a large number of generally less common alleles were also identified. Allele *CYP2D6*2* was significantly over-represented in South Central Asians (0.295) and Europeans (0.277), while *CYP2D6*35* was significantly over-represented in Europeans (0.055, Table S2). Alleles *CYP2D6*17, *29*, and *\*45* were significantly over-represented in African populations (Sub-Saharan African and African Americans/Afro-Caribbean) and under-represented in other populations.

## DISCUSSION

More than a year after the outbreak of the SARS-CoV-2 virus worldwide, very few drugs such as remdesivir have been proven to be effective against COVID 19^26^. In the absence of effective drugs, and until herd immunity is achieved globally, the risk of new variant outbreaks and mortality remain high. Having diverse population genetic information and genetic databases, could help clinicians avoid additional complications while treating COVID-19 patients. This work provides evidence for two very important drugs for which therapeutic actions are available based on level 1 evidence involving pharmacogenetic biomarkers and clinical pharmacogenetic guidelines. Such biomarkers were extensively studied and had already been validated in clinical practice^25^. Several genetic markers were analyzed for Jordanian Arab in context of diverse ethnic backgrounds to identify population differences in responses and toxicity events associated with atazanavir and fluvoxamine. Results from this study showed that pharmacogenomics studies can be leveraged to enhance the understanding of adverse reactions to the treatment of COVID-19 symptoms and support advancement of drug development pipelines.

MDS analysis showed that the Jordanian Arab population clustered within European and Near Eastern populations (Fig. 1). MDS results were further validated by the pairwise Fst values. These analyses are in agreement with recent studies using large-scale genomics that appeared to have facilitated and maintained admixture between culturally different populations^27^. In general, the Jordanian population was not significantly different from their Levantine neighbors, and fit consistently into a Middle East-Anatolia-Balkan-Caucasus geographic and genetic continuum^28^.

Of the 101 individuals of Jordanian Arab descent, ∼15% were poor metabolizers of atazanavir based as the first sequence variant analysis of *UGT1A1* for a Near Eastern population. However, mapped frequencies of *CYP2D6* and *UGT1A1* genotypes based on both the PharmGKB meta-analysis^25^, and the CPIC updated reports^15,24^, showed that East Asian populations are approximately 6.7x more likely to show impaired fluvoxamine metabolism than other groups and that African populations are approximately 7x more likely to show impaired atazanavir metabolism than East Asian populations (Fig. 3, Table S2). Therefore, a higher proportion of European, South Central Asian, Near Eastern, American, Latino, Oceanian ancestry populations have normal fluvoxamine or atazanavir metabolism, and therefore are less susceptible to complications for these two drugs.

**Figure 3.**
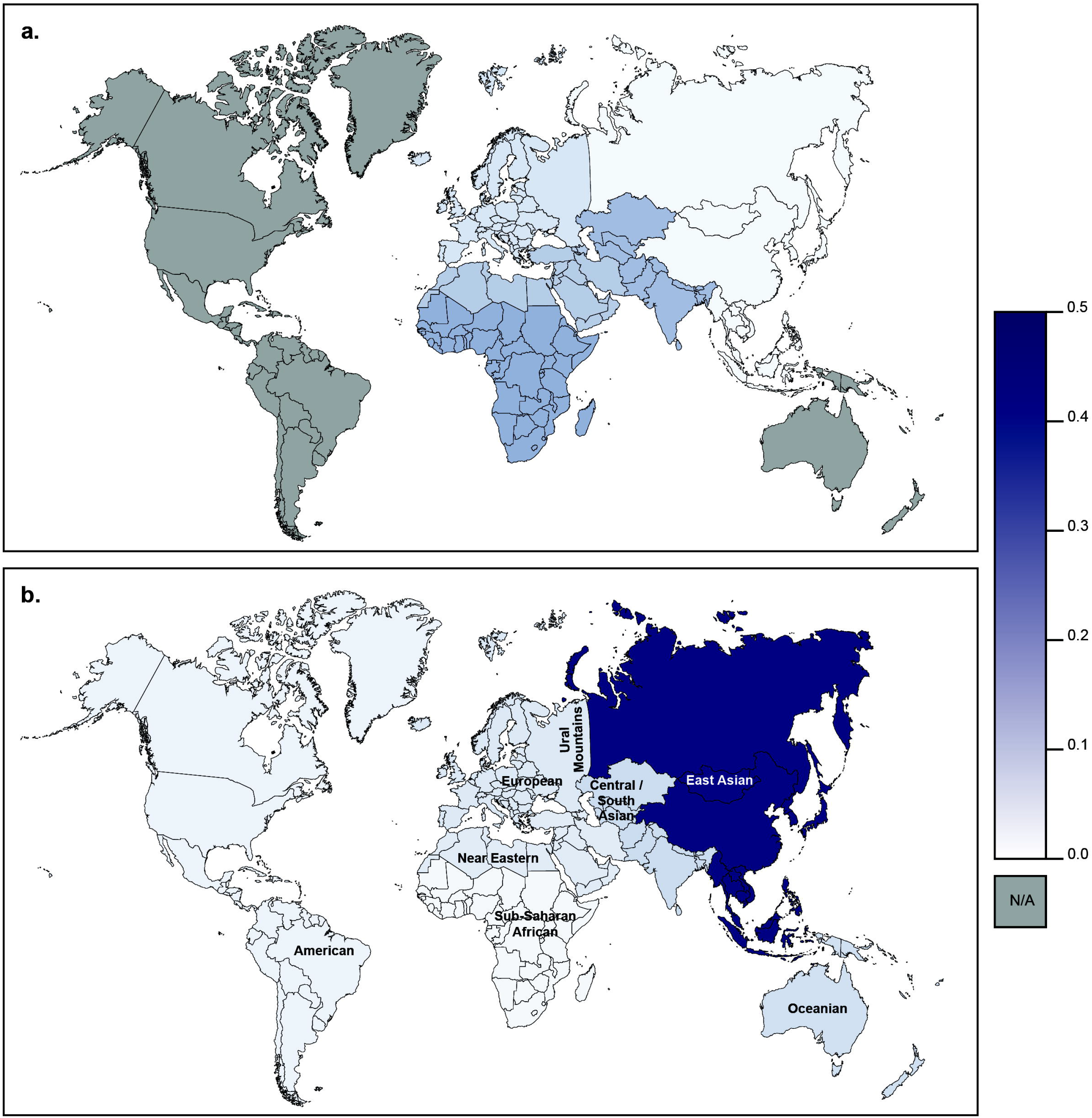
Global frequency map of variability in level 1A alleles and impact on atazanavir and fluvoxamine response.

Although our simulation is based upon phenotype allele frequencies instead of directly assaying patient biospecimens, these allele frequencies were previously well-described, and were already validated in clinical practice^29^. The adjusted expected allele frequencies based on the racial demographics of our cohort and expected population allele frequencies are dependent on population ancestry. Although precision medicine should be considered in COVID-19 treatment, there are still many challenges to face. Firstly, a suitable strategy for treatment of COVID-19 through integrating pharmacogenomics data is still lacking. A well-designed gene detection panel is important to fully characterize patient genotypes in relation to the drug effects^30^. Secondly, the best treatment regime can be optimized for each individual patient, where both the efficacy and the safety can be guaranteed. Additional research is necessary to clarify whether any other variants should be incorporated into clinical decision making.

Existing *CYP2D6* and *UGT1A1* genotype results may provide the potential benefit of identifying populations who are at an increased risk of experiencing adverse drug reactions or therapeutic failure. Collectively, This work demonstrates the capability and application of large-scale pharmacogenomics studies to elucidate genetic variation effects on treatment efficacy in COVID-19 patients. Ultimately, the implementation of pharmacogenetics in clinical settings is increasing as it leads to more efficient and cost-effective treatments. In times of a global healthcare crisis, these therapeutic improvements become crucial.

## METHODS

### Sample collection

This study retrospectively included 101 unrelated Jordanian participants, of which 56 were male and 45 were female. After a signed informed consent, a 3 ml venous blood sample was collected in 3 ml EDTA tubes from each participant at the Princess Haya Biotechnology Centre between May 2010 and December 2011. Blood samples were stored at 4°C until DNA extraction. Ethics approvals was provided by the Institutional Review Board (IRB) of the Jordan University of Science and Technology (registration number 67/2/2013), and performed in accordance with the principles enshrined in the Declaration of Helsinki.

### DNA extraction and genotyping

Genomic DNA was extracted from each blood sample using the QIAamp DNA Micro Kit (Qiagen, Hilden, Germany) according to the manufacturer’s instructions. The quality of the purified DNA was determined by using a NanoDrop ND-1000 spectrophotometer (Thermo Fisher Scientific, Waltham, MA USA).

Genotyping was done using the Affymetrix DMET (Drug Metabolizing Enzymes and Transporters) Plus Premier microarray assay (Santa Clara, CA, USA) to test for drug metabolism associations. The DMET array contains 1,936 drug metabolism markers consisting of 1,931 single nucleotide polymorphisms (SNPs) and five copy number variations (CNVs) in 225 genes, including 47 phase I enzymes, 80 phase II enzymes, 52 transporters and 46 other genes. These genetic variants were multiplex genotyped using molecular inversion probe (MIP) technology^31^. The profiles for the genotyping call rates and concordance comparisons, were generated by the DMET console software v1.3 (Thermo Fisher Scientific, Inc., Waltham, MA, USA), based on the Bayesian robust linear model with Mahalanobis (BRLMM) distance classifier algorithm^32,33^. Genotypes were determined for each SNP site and reported as homozygous wild-type, heterozygous, homozygous variant, and ‘no call’ where no genotype was called. Genotype call rates ranged from 99% to 100%. SNPs with a call rate of less than 99% were excluded from subsequent analyses. Statistical and genetic analyses were performed for selection and validation using Microsoft Excel and SPSS v16 (Fig. 4). The genotype and allele frequencies were calculated and tested for deviations from Hardy-Weinberg equilibrium using the chi-square (χ2) test (*p*L>L0.05). Fisher’s exact test was used to correct for small population size. These were implemented with the *HWChisq* and *HWextact* functions in HardyWeinberg R package^34^. A Holm’s sequential correction for multiple comparisons was applied^35^.

**Figure 4.**
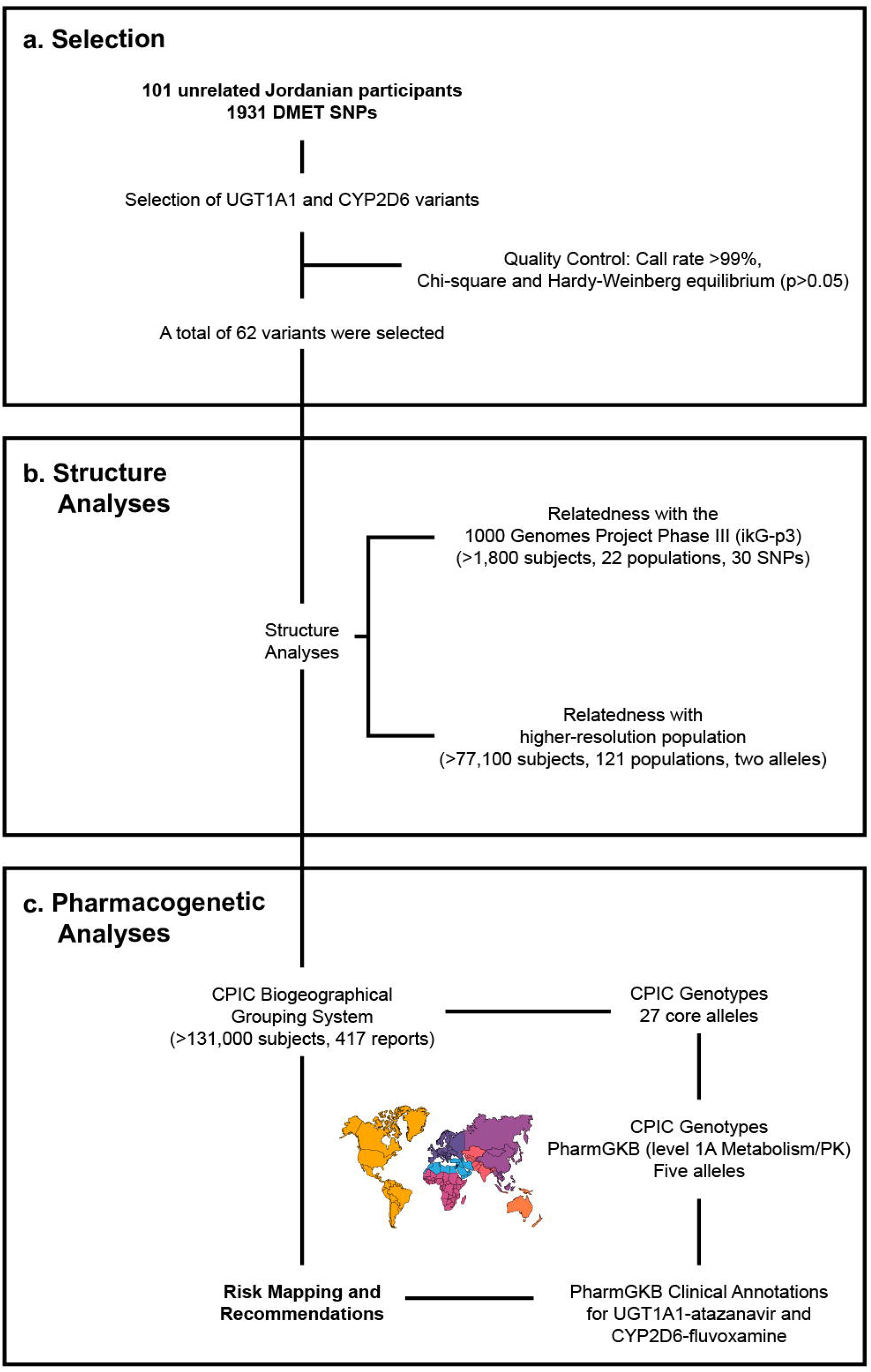
Flow chart depicting various steps involved in variant selection, analysis, and annotation. (**a**) Selection of *UGT1A1* and *CYPT2D6* variants. (**b**) Population structure analysis between the Jordanian Arab population and three defined datasets. (**c**) Pharmacogenetics analyses with nine geographically-defined groups compiled with CPIC guidelines (>100,000 subjects, 417 reports) estimating genotype-predicted phenotype status across world populations.

### Population structure analyses

Population structure was examined using Wright’s Fixation Index (Fst) for pairwise distances between populations based on the *UGT1A1* (12 SNPs) and *CYP2D6* (18 SNPs) data to identify the ancestral relatedness between the Jordanian Arabs and other populations. Two defined datasets were considered:

Ethnically defined populations by 1kG-p3 dataset, consisting of 1810 individuals from 22 populations within four defined ancestral groups (Table S3). The admixed populations were excluded to simplify the ethnic identification analyses.

1. African (AFR): Americans of African Ancestry in SW, USA (ASW), Esan in Nigeria (ESN), Gambian in Western Divisions in the Gambia (GWD), Mende in Sierra Leone (MSL), Luhya in Webuye, Kenya (LWK), Yoruba in Ibadan, Nigeria (YRI) and African Caribbeans in Barbados (ACB).
2. European (EUR): Utah Residents (CEPH) with Northern and Western European Ancestry (CEU), Finnish in Finland (FIN), British in England and Scotland (GBR), Iberian Population in Spain (IBS) and Toscani in Italia (TSI).
3. East Asian (EAS): Han Chinese in Beijing, China (CHB), Chinese Dai in Xishuangbanna, China (CDX), Southern Han Chinese (CHS), Japanese in Tokyo, Japan (JPT) and Kinh in Ho Chi Minh City, Vietnam (KHV).
4. South Asian (SAS): Bengali from Bangladesh (BEB), Indian Telugu from the UK (ITU), Punjabi from Lahore, Pakistan (PJL), Tamil from the UK (STU) and Gujarati Indian from Houston, Texas (GIH).

One more higher-resolution groups of populations consisting of 77,176 individuals from 121 global reports from the CPIC updated report in October 2019^24^. Divergences were visualized by multidimensional scaling analysis (MDS) using *the metaMDS function* in the vegan R package (Table S4)^36^. Fixation indices were estimated with the *calcFst* function in the polysat R package^37^.

### Pharmacogenetic analyses

The frequencies of the 27 actionable pharmacogenetics biomarkers were assessed cumulatively for the Jordanian Arabs against nine biogeographical groups, consisting of 130,856 individuals from 417 global populations^15,24^.

These nine groups were defined by global autosomal genetic structure and based on data from large-scale sequencing initiatives and are used to illustrate the broad diversity of global allele frequencies in this study. Furthermore, this biogeographic grouping system meets a key need in pharmacogenetics research by enabling consistent communication of the scale of variability in global allele frequencies and are now used by PharmGKB and CPIC. The genotypic data of individuals from 417 global populations were downloaded from the CPIC updated reports.

1. American (AME): The American genetic ancestry group includes populations from both North and South America with ancestors predating European colonization, including American Indian, Alaska Native, First Nations, Inuit, and Métis in Canada, and Indigenous peoples of Central and South America.
2. Central/South Asian (SAS): The Central and South Asian genetic ancestry group includes populations from Pakistan, Sri Lanka, Bangladesh, India, and ranges from Afghanistan to the western border of China.
3. East Asian (EAS): The East Asian genetic ancestry group includes populations from Japan, Korea, and China, and stretches from mainland Southeast Asia through the islands of Southeast Asia. In addition, it includes portions of central Asia and Russia east of the Ural Mountains.
4. European (EUR): The European genetic ancestry group includes populations of primarily European descent, including European Americans. We define the European region as extending west from the Ural Mountains and south to the Turkish and Bulgarian border.
5. Near Eastern (NEA): The Near Eastern genetic ancestry group encompasses populations from northern Africa, the Middle East, and the Caucasus. It includes Turkey and African nations north of the Saharan Desert.
6. Oceanian (OCE): The Oceanian genetic ancestry group includes pre-colonial populations of the Pacific Islands, including Hawaii, Australia, and Papua New Guinea.
7. Sub-Saharan African (SSA): The Sub-Saharan African genetic ancestry group includes individuals from all regions in Sub-Saharan Africa, including Madagascar.
8. African American/Afro-Caribbean (AAC): Individuals in the African American/Afro-Caribbean genetic ancestry group reflect the extensive admixture between African, European, and Indigenous ancestries and, as such, display a unique genetic profile compared to individuals from each of those lineages alone. Examples within this cluster include the Coriell Institute’s African Caribbean in Barbados (ACB) population, the African Americans from the Southwest US (ASW) population, and individuals from Jamaica and the US Virgin Islands.
9. Latino (LAT): The Latino genetic ancestry group is not defined by an exclusive geographic region, but includes individuals of Mestizo descent, individuals from Latin America, and self-identified Latino individuals in the United States. Like the African American/Afro-Caribbean group, the admixture in this population creates a unique genetic pattern compared to any of the discrete geographic regions, with individuals reflecting mixed native and indigenous American, European, and African ancestry.

The total frequency of six SNPs with a Level 1A for the Jordanian Arab population within nine geographically-defined groups were mapped for global impact visualization of allele frequency on atazanavir and fluvoxamine response (Table S2, Fig. 4). Inferred frequencies for *UGT1A1*1* and *CYP2D6*1* were excluded from our biogeographical analyses as no population studies have tested for all known variant alleles, and *\*1* was not genotyped directly in many studies^15,24^.

## Data Availability

The datasets generated during the current study are included in the supplementary files. Publicly available repositories used in the analyses were previously published under PMID: 26417955 and 25974703.

https://api.pharmgkb.org/v1/download/file/attachment/UGT1A1_frequency_table.xlsx

https://api.pharmgkb.org/v1/download/file/attachment/CYP2D6_frequency_table.xlsx

## DECLARATIONS

### Ethics approval and consent to participate

Signed informed consent was obtained from all participants recruited for this study under the ethical approval from University Review Committee for Research on Humans at Jordan University of Science and Technology (JUST) registered under [67/2/2013] on 4/7/2013, and performed in accordance with the principles enshrined in the Declaration of Helsinki.

### Consent for publication

Consent for publication of the data obtained in this study was retrospectively approved, following de-identifying information of individuals.

### Availability of data and materials

The datasets generated during the current study are included in the supplementary files. Publicly available repositories used in the analyses were previously published under PMID: 26417955 and 25974703,and downloaded from https://api.pharmgkb.org/v1/download/file/attachment/UGT1A1_frequency_table.xlsx, and https://api.pharmgkb.org/v1/download/file/attachment/CYP2D6_frequency_table.xlsx.

## Authors’ contributions

The author conceived the research study, performed all the analyses reported and wrote the manuscript.

## Acknowledgements

The technical staff from Princess Haya Biotechnology Center at the Jordan University of Science and Technology provided their assistance with this study.

